# Gestational Environment Captured by the Neonatal Metabolome is not Predictive of Later Inflammatory Bowel Disease

**DOI:** 10.64898/2026.02.18.26346468

**Authors:** Alice Fracchia, Jonas J. Rudbaek, Kaustubh Chakradeo, Tine Jess, Filip Ottosson, Aleksejs Sazonovs

**Author notes:** **Correspondence** Aleksejs Sazonovs, A.C. Meyers Vænge 15, 2450 Copenhagen SV, Denmark.

## Abstract

**Background:** Gestational exposures may contribute to the newborn’s lifetime risk of inflammatory bowel disease (IBD). While gestational influences are associated with IBD onset, the causality and confounding of such exposures are difficult to ascertain. The neonatal metabolome provides a metabolic snapshot of gestational influences.

**Objective:** We tested the neonatal metabolome’s ability to predict future IBD, to assess whether gestational exposures are reflected in early molecular precursors of the disease.

**Methods:** We profiled dried blood spots from 520 newborns who later developed IBD and matched controls using high-resolution untargeted mass spectrometry metabolomics (1,350 QC-passing metabolites). Genotyping was available for 1,009 of these individuals. PERMANOVA confirmed assay sensitivity to gestational exposures, gradient boosting was used for prediction.

**Results:** The neonatal metabolome significantly captured maternal smoking, birth weight, and gestational age (p < 0.001), but explained minimal variance in IBD status (R^2^ = 0.09%, p = 0.390) and showed no predictive power for IBD (AUC = 0.51, 95% CI 0.50–0.52, p = 0.585). Stratifying by disease subtype and age of onset did not improve performance. In contrast, genetic risk scores were modestly predictive (CD: AUC = 0.64, p < 5.11×10⁻^14^; UC: AUC = 0.63, p < 7.65×10⁻¹²), but uncorrelated with neonatal metabolomic profiles (CD: p = 0.650; UC: p = 0.970), suggesting a later-age effect.

**Conclusions:** Using a large, comprehensively profiled cohort, we demonstrate that neonatal metabolomic profiles sensitively capture gestational signatures, but not the overall future IBD risk. Our findings suggest that most IBD risk accumulates later in life, beyond gestational molecular imprints.

## Introduction

Inflammatory bowel disease (IBD) is a chronic inflammatory condition with a complex aetiology, driven by the interplay of genetic susceptibility and environmental exposures^1–4^. Genetic factors are strongly linked to the disease but remain poor predictors of its onset^5–7^. Heritability estimates suggest that the common genetic variation accounts for around 42% and 25% of disease susceptibility for Crohn’s disease (CD) and ulcerative colitis (UC), respectively^8^. This “genetic ceiling”, coupled with the rapidly rising incidence of IBD^9^, underscores the important role of environmental factors in disease development. Such exposures can modulate disease risk and may be especially influential during critical periods of immune system development, like the early life^1,10,11^.

Gestational age, delivery mode, maternal smoking, and diet during pregnancy are some of the established factors influencing the neonatal molecular state^12–18^, with potential long-lasting consequences for subsequent immune and metabolic development. Variation in the early molecular landscape has been associated with paediatric conditions, such as kidney disorders, respiratory illnesses, and necrotizing enterocolitis, as well as later onset disorders, e.g., cardiometabolic factors and immune-mediated disease, such as type 1 diabetes and hyperthyroidism^19–24^.

Recent studies have therefore focused on the role of gestational exposures in shaping long-term immune function and IBD susceptibility^25–27^. Disentangling the effect of these prenatal exposures from postnatal or socioeconomic confounders remains a major challenge in observational epidemiology. While individual environmental factors are often difficult to isolate, untargeted metabolomics helps to address this challenge by quantifying their downstream biological effects.

The metabolomic profile combines the small-molecule metabolites, including amino acids, lipids, and small peptides, circulating within a biological system. It reflects the downstream outcome of gene expression, protein activity, and environmental interaction, offering a dynamic snapshot of the physiological state at a given time^28,29^. By profiling metabolites around the time of birth with a broad, untargeted assay, we can access an integrated view of maternal influences and foetal metabolism prior to postnatal confounders. Although technical variation can influence metabolite detection, this approach offers broad coverage of the metabolome, enabling exploration beyond a narrow set of predefined targets^29^.

Previous research has investigated associations between neonatal metabolomic profiles and IBD, identifying links between levels of certain amino acids, nucleotides, peptides, and acylcarnitine and future IBD onset^30^. A recent study highlighted the absence of predictive value of a targeted set of routine and narrowly selected neonatal disease-screening metabolites for paediatric IBD^31^. The goal of this study was to evaluate whether gestational exposures, as captured by the neonatal metabolomic profile at birth, are predictive of the future IBD risk.

## Materials and methods

### Birth-cohort design and case–control selection

Using a nested case-control framework, we identified IBD cases as individuals having at least two hospital contacts, within two years^9^, registered in the Danish National Patient Registry. Diagnoses are encoded through the ICD-8 (56301, 56302, 56308, 56309, 56319, 56904) and ICD-10 codes (DK50, DK51). Controls were matched on gestational age, birth weight, sex, and sample collection day^30^. The cohort was designed to include a similar number of paediatric-(n = 222, before 19 years) and adult-onset (n = 298, after 19 years) patients.

### DBS collection, storage, and metabolomics workflow

Dried Blood Spots (DBS) have been routinely collected in Denmark since 1981 for neonatal screening purposes (sampling day distribution in Fig. S1B, Supplementary Materials). The samples are preserved on Guthrie cards at -20°C within the Danish National Biobank, serving as a standardised longitudinal resource for metabolic research^32^. Mass spectrometry analysis was performed on a timsTOF Pro mass spectrometer coupled to an ultra-high-performance liquid chromatography Elute system, Bruker Daltonics (Billerica, MA, USA). Details about sample preparation and mass spectrometry analysis have been described previously^30^.

### Metabolites pre-processing

A total of 10,269 metabolites were measured. Metabolomics pre-processing was performed in MZmine^33,34^, and only features with less than 25% missing values (n = 5133) were retained for quality control. Metabolites passing quality control were imputed using the limit-of-detection method, resulting in 1350 reliably measured metabolites. Inter- and intra-batch effects were corrected using WaveICA^35^. Data were subsequently z-score normalised.

### Phenotypic data sources

Gestational, perinatal, clinical, and demographic factors were obtained from the Danish National Patient Register, the National Birth Register, and the Central Person Register through the Danish unique personal identifier, which enables linking information across records. Variables included in the analysis were: postnatal sampling day, sex, delivery mode, gestational age, year of birth, birth weight, maternal smoking (either anytime during pregnancy or during the last trimester), maternal BMI, maternal IBD, paternal IBD, future IBD onset, and year of IBD onset. The missing postnatal sampling day was imputed as the mean sampling day of the cohort (6 days) for 71 individuals. Gestational age for 16 individuals was imputed with predictive mean matching. The overall maternal smoking status during pregnancy was unknown for 8 individuals (0.77% never replied), while information about smoking during the last trimester was missing for 234 individuals (22.5%). Birth weight was missing for 444 individuals (42.3%).

### Additional molecular measurements

#### Genotype data

Genotype data were available for 1009 (97%) individuals in the cohort (504 cases and 505 controls). DNA was extracted and genotyped using a modified version of the Global Screening Array v3 from Illumina (San Diego, CA, USA). Variants with a minor allele frequency less than 0.01 were excluded. Quality Control (QC) followed the protocol described in a previous publication^36^. After QC, genotype imputation was performed using the 1000 Genomes Project reference panel. GRS weights calculated via LDPred2 by *Middha et al.*^5^, based on the *de Lange et al.*^6^ summary statistics, were applied for our cohort.

#### Cytokine data

The concentration of 6 cytokines (IFN-γ, TNF-α, IL-1β, IL-4, IL-6, and IL-17A) was measured at birth and available for 946 individuals, of which 365 (35%) overlapped with the metabolome cohort. The measurements were performed with the S-PLEX proinflammatory panel 1 kit (#K15396S, MesoScale Diagnostics, USA), and missing values due to the low limit of detection were replaced by half the lowest detectable value^25^.

### Variance analysis (UMAP, PERMANOVA, and Lasso)

To investigate the totality of the measured metabolites in a low-dimensional space, we used a non-linear dimensionality reduction method, UMAP^37^. To quantify the fraction of variance in the overall metabolomic profile explained by each of the maternal factors, we applied PERMANOVA (Permutation Analysis of Variance)^38^ - a non-parametric multivariate test which assesses whether group centroids differ significantly based on an Euclidean distance matrix. It returns a pseudo F-statistic and a corresponding pseudo-R^2^, which quantifies the proportion of total variance explained by group differences. In addition, Lasso regression^39^ was used to evaluate the fraction of variance in the variable explained by the combination of metabolites. This adjusted regression allowed for assessing the association between an individual outcome and a multivariate predictor (Supplementary Materials for more details).

### Predictive modelling

The data were randomly split between training and test sets (70:30 ratio), preserving the matching between cases and controls. A broad set of prediction models (Lasso, Random Forest, Support Vector Machine, Gradient Boosting, eXtreme Gradient Boosting [XGBoost]) was compared to assess the predictive power of metabolite profiles for IBD onset. Model hyperparameters were tuned through a 5-fold cross-validated grid search on the training data (Tab S1, Supplementary Materials). The best-performing model was evaluated on the test set with AUC as the performance metric for binary classification, and C-index for regression.

To obtain a reliable estimate of the predictive performance, the model was run 30 times with different random seeds for the train-test split, and performance metrics were averaged across runs. The optimal number of runs was chosen as the smallest number of runs after which the average AUC converged. For continuous outcomes, the C-index is used to measure the performance (Supplementary Materials for additional details). For binary variables with unbalanced distribution between classes, such as delivery mode, smoking status, or parental IBD, we downsampled the majority class to match the minority class size. Confidence intervals were calculated with 1000 bootstraps for both continuous and binary variables. Permutation p-values were calculated with 1000 permutations. The significance threshold was adjusted with Bonferroni correction for the number of multiple tests.

## Results

### Cohort characteristics and quality control

The cohort included a total of 1,040 individuals: 520 newborns who later developed IBD (276 CD and 244 UC) and 520 matched controls. Age at onset among cases ranged from 0 to 28 years, with approximately half of the cohort being diagnosed before the age of 19. Sex was evenly represented in the cohort (53% female). Demographic and clinical characteristics are summarised in Table S2 in the Supplementary Materials. Details about the distribution of age of onset and postnatal sampling day are reported in Fig. S1 of the Supplementary Materials.

The first 600 principal components (PCs) derived from metabolites explained around 99% of the total variance in the dataset. None of the first 30 components (60% of the total variance) had evidence of association with IBD case-control status (p > 0.100). UMAP built from 600 PCs did not reveal any clustering by IBD status (Fig. 1A) and was primarily driven by differences in sampling year and the postnatal sampling day relative to birth. Birth year and postnatal sampling day emerged as the strongest factors, accounting for 3.7 and 2.2% of the total variance (Figure S2, Supplementary Materials). To avoid potential confounding and to concentrate on the gestational rather than sampling effects, both factors were regressed out of the metabolite profiles before downstream analysis.

**Figure 1:**
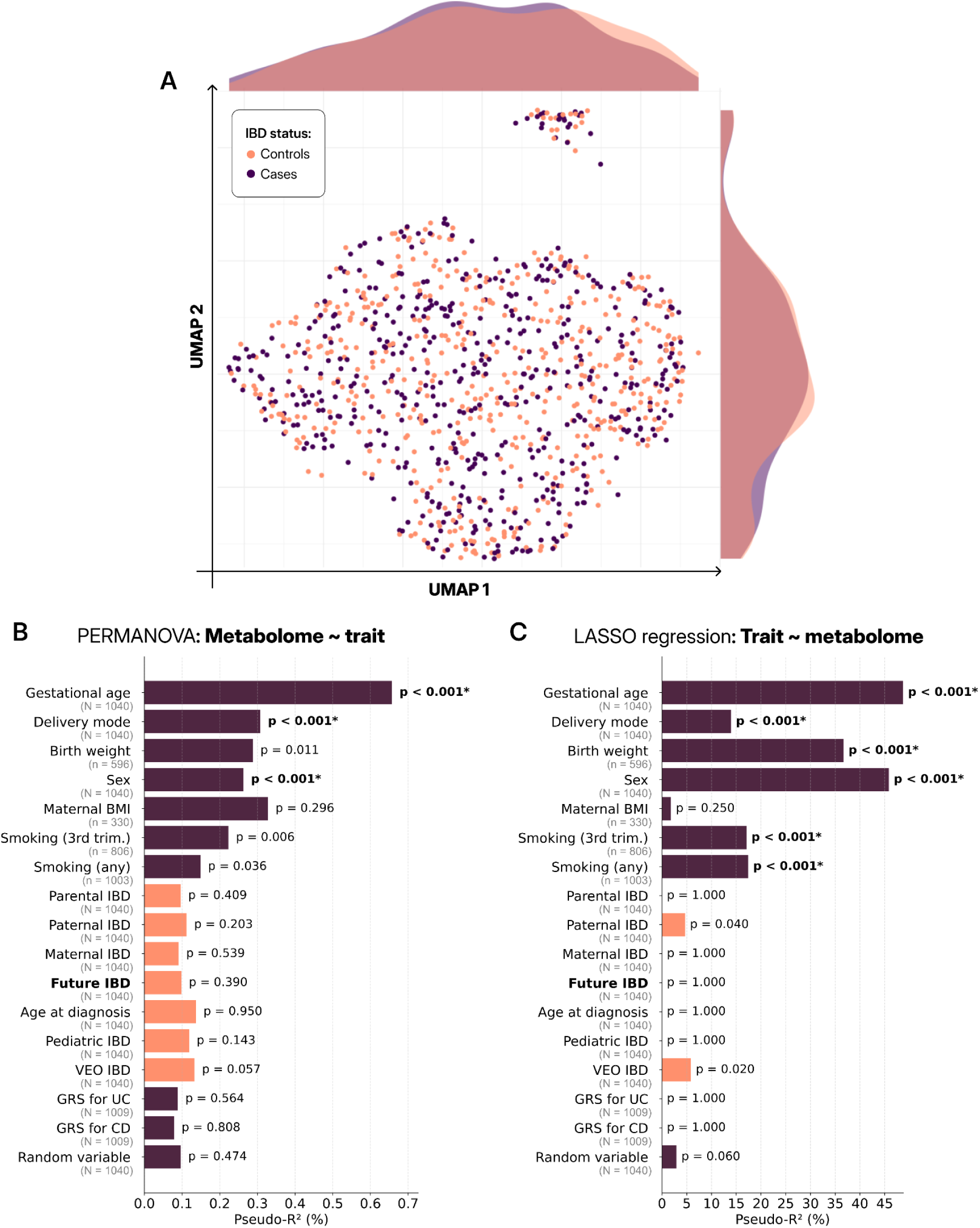
(A) UMAP representation of the 600th first PCs of the metabolite profiles, color-coded according to case-control status. There is no noticeable difference in density distributions between the two statuses. The small cluster does not associate with any particular variable. (B) The contribution in percentage to the total metabolome variance for different perinatal, maternal, and genetic factors. Gestational age, delivery mode, and sex are the only statistically significant contributions, explaining respectively 0.66, 0.31, and 0.25% of the total variance. Pseudo- ^2^ was calculated with PERMANOVA. (C) The variance in maternal factors as captured by the metabolome. 49%, 46%, 37%, and 14% of the variance in gestational age, sex, birth weight, and delivery mode can be significantly explained by a difference in the metabolite profiles. Smoking is also well captured through variation of the metabolome. Significant p-values are indicated with an asterisk. IBD-related variables are represented in orange, with future IBD risk in bold.

### Factors influencing the neonatal metabolome

Before evaluating IBD prediction, we first confirmed that neonatal metabolomics in DBS was sensitive to known gestational and perinatal determinants. We assessed the contribution of maternal environmental factors and other variables to variation in the neonatal metabolome using PERMANOVA with an Euclidean distance matrix. Among biological factors, gestational age, delivery mode, and sex were the only variables to show statistically significant contributions to the variance in the metabolome (Fig. 1B). In contrast, maternal factors (maternal smoking, IBD, and BMI), perinatal traits (birth weight), genetic predisposition (parental IBD and genetic risk scores), and future IBD status contributed negligibly. IBD status explained negligible variance, comparable to a random binary label, suggesting that early-life metabolic profiles are not strongly indicative of future disease onset. Despite their limited impact on overall metabolomic variance, Lasso regression revealed that maternal smoking and birth weight impart discrete, statistically significant alterations to the overall neonatal molecular landscape (Fig. 1C).

### Parental IBD: minimal metabolomic imprint

We assessed the variance in the neonatal metabolome explained by maternal and paternal IBD status. Paternal IBD was included as a proxy for inherited genetic risk, since it does not involve direct in utero exposure, unlike maternal IBD. PERMANOVA results indicated that neither of the parental IBD statuses accounts for a significant portion of variance, each explaining less than around 0.1% (p > 0.200) (Figure 1), indicating a weak contribution of inherited genetic IBD effects to the neonatal metabolome.

### Predictive performance of the neonatal metabolome

We applied a binary classification model to evaluate the predictive power of the neonatal metabolome on future IBD onset. We included other maternal and perinatal factors as benchmark traits with known metabolic associations, serving as positive controls for model performance (Fig. 2A). Sex and gestational age yielded the best results, with AUCs over 0.65. We found no evidence of predictive performance beyond chance (95% CI 0.50–0.52, p = 0.585). Stratifying by disease subtype and age of onset did not improve the predictive power (Fig. 2B).

**Figure 2:**
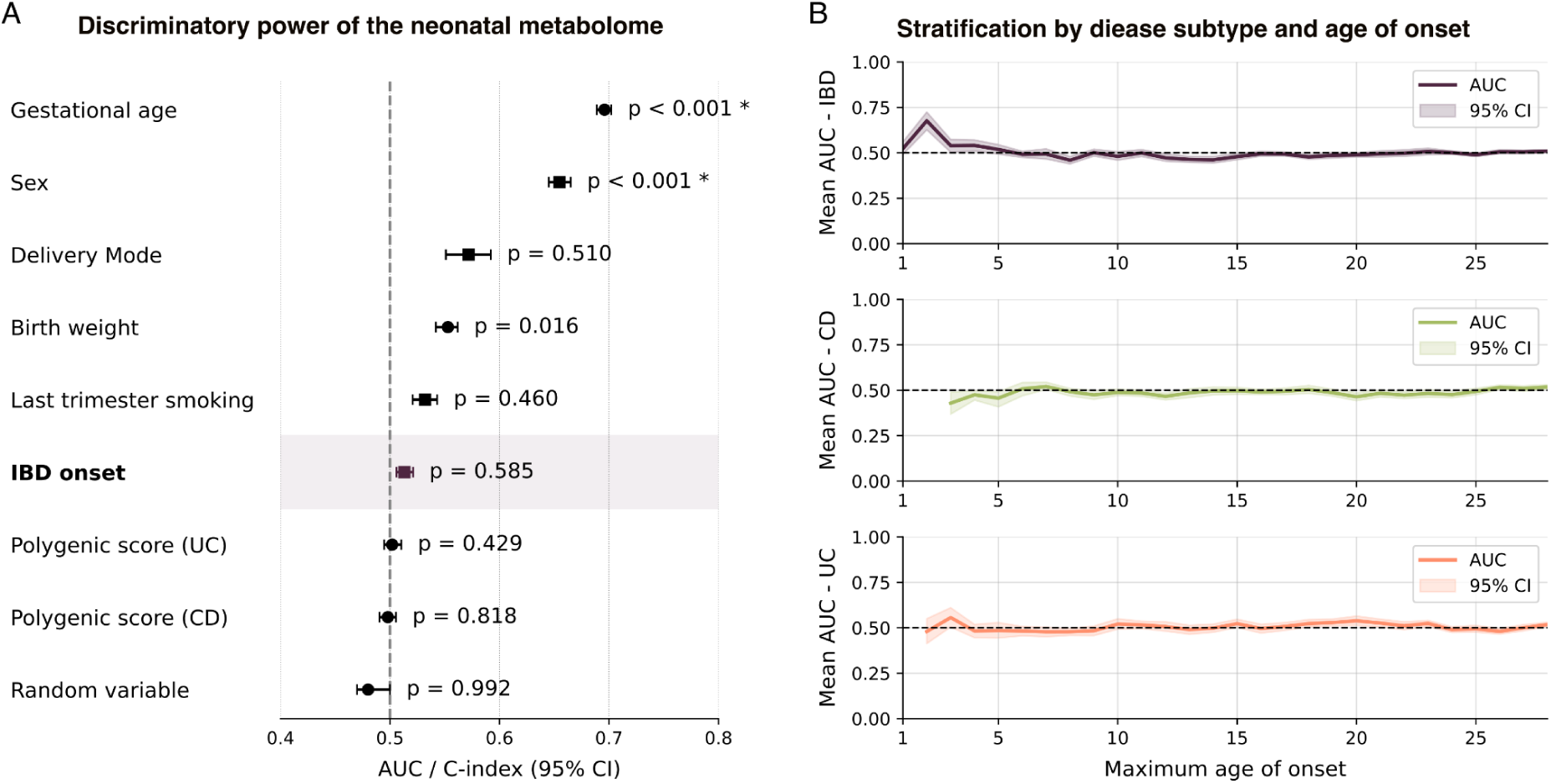
(A) Predictive performance of XGBoost in terms of AUC and C-index for, respectively, binary and continuous outcomes. Among binary traits, sex could be predicted with significant accuracy (AUC = 0.65), whereas parental or future IBD status could not. For continuous traits, gestational age can be significantly predicted with decent accuracy, contrary to genetic risk scores. Error bars indicate 95% CIs, and p-values were determined through permutation testing for 1000 iterations. Asterisks indicate statistical significance after Bonferroni correction. (B) IBD onset prediction stratified by disease subtype and onset age. The trend of no predictive power is stable across runs, and slightly higher AUCs are compromised by a very low sample size for very-early onset. Each point was generated by running the model 30 times, averaging the AUCs, and computing the bootstrapped confidence intervals. Years 1-2 and 1 for CD and UC, respectively, were excluded due to insufficient sample size to run the model.

### Additional molecular measurements: genotyping and cytokine dataset

To explore whether other early-life molecular layers could contribute to IBD prediction, we applied the same predictive framework to available genotyping and cytokine datasets, and assessed whether genetic risk scores were reflected in the neonatal metabolome. While genetic risk scores were modestly predictive of future IBD onset (AUC = 0.64, p < 5.11×10⁻^14^ for CD; AUC = 0.63, p < 7.65×10⁻¹² for UC), the performance of the cytokines was consistent with the limited signal observed using metabolomic profiles, resulting in a model with an AUC of 0.56 (95% CI 0.55-0.58, p = 0.50). Additionally, genetic risk scores were uncorrelated with neonatal metabolomic profiles (p = 0.970 for UC, p = 0.650 for CD), suggesting later-life action of the genetic effects predisposing to IBD.

## Discussion

Untargeted metabolomics offers a comprehensive view of an individual’s circulating metabolites, small molecules produced by their metabolism, providing a snapshot of their physiological state. Profiling neonatal blood samples collected shortly after birth captures the influence of gestational factors – such as delivery mode, gestational age, and diet – on the newborn’s metabolomic profile^13,40,41^. In adults, previous research has reported associations between individual metabolites and incident IBD^42–44^. Our prior analysis of this cohort linked 25 metabolites, measured in individuals at birth, to future IBD onset^30^. Here, we examined the predictive capacity of neonatal metabolomic profiles and quantified the relative contribution of this early-life molecular signature to IBD risk.

Based on this large and representative cohort, our findings indicate that these individual signals and environmental imprints do not translate into a predictive model. The combined predictive effect of the gestational factors it reflects is near zero. The same at-birth metabolomic layer robustly captures gestational and perinatal exposures (positive controls), yet carries little information about later IBD case–control status. This pattern is most consistent with IBD-relevant molecular divergence emerging primarily postnatally.

While perinatal factors leave discernible metabolic signatures at birth, their overall impact on long-term IBD susceptibility appears minimal. This is not contradictory: effects can be statistically detectable yet too small to yield meaningful individual-level discrimination. Despite minimal predictive value, the appeal of these signals is aetiological: measured before most later-life cumulative effects, they provide a cleaner view of background biology that is harder to disentangle in later-life samples alone.

Genetic risk scores demonstrate a significant, although modest, predictive value for both Crohn’s disease and ulcerative colitis. Notably, we found no significant correlation between genetic risk and neonatal metabolomic profiles for either condition. This contrasts with later-life pre-diagnostic profiling, where measures from dynamic omics are partially collinear with genetic risk scores^42^. This lack of correlation suggests that the inherent IBD risk either accumulates over time, and thus is too weak for us to identify at birth, or that it manifests in non-genetic molecular signatures only after environmental exposures (e.g., *NOD2* variants are associated with higher IBD risk, plausibly through altered bacterial recognition^45^). Our findings suggest that the majority of IBD risk accumulates later in life and is not substantially driven by gestational and perinatal factors that impact the early-life metabolomic profile.

While no predictive value was found for either paediatric or adult-onset IBD, we cannot rule out a signal in very early-onset patients, who were rare in our cohort. Limitations include the contribution of storage year and sampling day relative to birth to variance in the metabolome, consistent with molecular degradation and the dynamic nature of the metabolome^32,46^. While our metabolomics protocol is broad, comprehensive, and untargeted, we do not capture all types of circulating metabolites. Expanding metabolite coverage by combining different measuring techniques could help address this limitation.

At-birth prediction of disease onset remains a desired avenue for common chronic conditions, including IBD. Of the three modalities assessed here (untargeted metabolomics, cytokines, and genetics), only genetic risk scores exhibited measurable predictive value for both CD and UC, albeit falling short of clinical relevance. While we cannot rule out additional predictive value of other dynamic omics in neonatal samples (e.g., methylation, transcriptomics, proteomics), broad metabolomics reflects neonatal physiology and gestational influences, and should therefore be sensitive to alterations occurring in those upstream layers.

In conclusion, our results show that neonatal metabolomes reflect selected perinatal characteristics but do not meaningfully predict future IBD risk. Genetic predisposition is present at birth, yet does not appear to be mirrored in the neonatal metabolic state. These results suggest that neonatal screening has limited utility for predicting IBD. Instead, future work should prioritise postnatal windows to better capture environmental risk exposures – while recognising a possible exception for very early-onset disease, which warrants targeted investigation.

In a large, population-based cohort, neonatal DBS metabolomics is highly sensitive to gestational and perinatal exposures but largely uninformative for later IBD prediction, suggesting that clinically relevant molecular signatures may emerge after birth.

## Data availability statement

The data from this study are protected by the Danish Data Protection Act and European Regulation 2016/679 of the European Parliament and of the Council (GDPR). Samples can be accessed through an application process at the Danish National Biobank. Requests for collaboration can be directed to the corresponding author. Analysis code is available on GitHub, linked in the Supplementary Materials.

## Ethics statements

### Patient consent

Patient consent was respected in accordance with the regulations set up by the Danish Data Protection Agency.

### Ethics approval

The study follows the ethical approval granted by the regional ethical committee of the Capital Region of Copenhagen (document number 06072022).

## Acknowledgements

This work was supported by the Danish National Research Foundation (grant DNRF148) and the Novo Nordisk Foundation (grant NNF23OC0087616).

