## Supplementary Material for "Gestational Environment Captured by the Neonatal Metabolome is not Predictive of Later Inflammatory Bowel Disease"

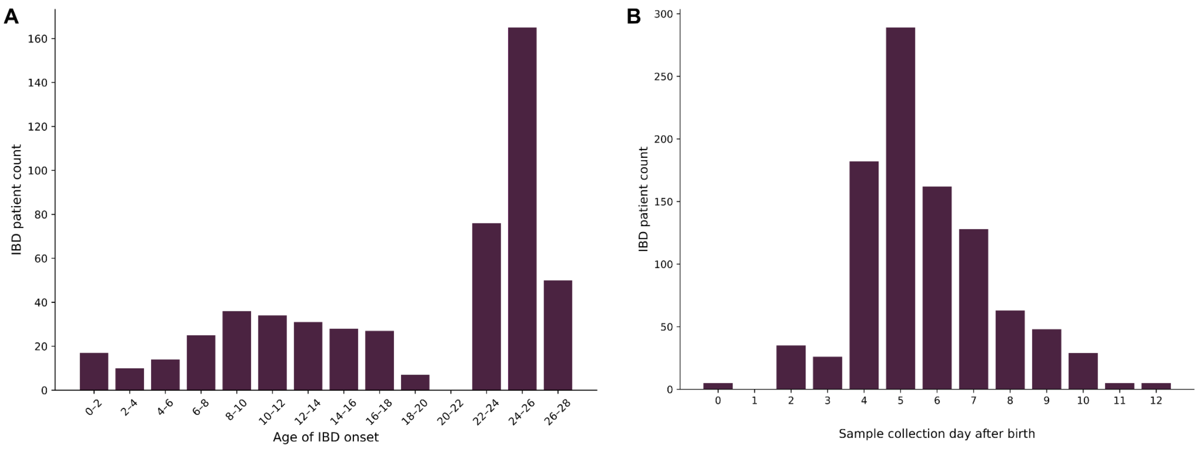


**Figure S1**: (A) Distribution of the age at IBD diagnosis in years for the cohort, which is representative of both pediatric and early-adulthood onset. (B) Distribution of the number of days after which samples were collected post-birth. Minimum displayed count is 5.

**Table S1**: Hyperparameter space for XGBoost tuning with 5-fold cross-validated gridsearch.

| Parameters | Purpose | Value 1 | Value 2 | Value 3 |
|---|---|---|---|---|
| Number of trees | Number of sequential decision trees in the ensemble that collectively form the final model. | 100 | 300 | 500 |
| Maximum tree depth | Controls the maximum depth of each tree, limiting model complexity and overfitting risk. | 3 | 5 | 7 |
| Learning rate | Scales each tree's contribution to the final prediction, trading training speed against generalisation. | 0.01 | 0.05 | 0.1 |
| Dataset subsample | Fraction of training samples randomly selected for each tree to reduce overfitting and variance. | 0.6 | 0.8 | 1.0 |
| Feature subsample | Fraction of features randomly selected for each tree or split to reduce feature correlation and overfitting. | 0.6 | 0.8 | 1.0 |

### C-index, Bootstrap CI, and Permutation p-value

*C-index* - Generalised AUC for continuous classification by measuring the probability that, for a randomly selected pair of samples, the predicted ordering matches the actual ordering of outcomes.

*Bootstrap CI* - For each bootstrap iteration, the metric values were randomly resampled with replacement, and the mean was computed. The 95% confidence interval was defined by the 2.5th and 97.5th percentiles of the resulting distribution of bootstrapped means.

*Permutation p-value* - To calculate a p-value, class labels were randomly permuted 1000 times without refitting the model. For each permutation, we computed the performance metric and compared it to the original metric. The p-value was defined as the proportion of permutation scores greater than or equal to the observed score.

**Table S2**: Demographic and clinical information of the cohort for both binary and continuous variables.

| Binary variables | Count (%) | Continuous variables | Mean (std, min-max) |
|---|---|---|---|
| Sex | | Birth year | 1997 (7, 1990-2018) years |
| Male | 492 (47%) | | |
| Female | 548 (53%) | | |
| Diagnosis | | Sampling age | 6 (2, 0-12) days |
| CD | 276 (53%) | | |
| UC | 244 (47%) | | |
| Delivery mode | | Gestational age | 280 (9, 224-305) days |
| Natural | 919 (88%) | | |
| C-section | 121 (12%) | | |
| Parental IBD | | Birth weight | 3512 (413, 1835-4800) g |
| Maternal | 47 (4.5%) | | |
| Paternal | 16 (1.5%) | | |
| Either | 62 (6%) | | |
| Any maternal smoking | | Age at diagnosis | 19 (8, 0-28) years |
| Yes | 267 (39%) | | |
| No | 765 (61%) | | |
| Maternal smoking - last trimester | | Maternal BMI | 25 (5, 17-62) |
| Yes | 168 (26%) | | |
| No | 638 (74%) | | |
| Onset type | | PRS for UC | -0.9 (1.0, -4.3-2.0) |
| VEO | 31 (3%) | PRS for CD | 1.1 (1.3, -2.8-4.9) |
| Pediatric | 211 (20%) | | |
| Adult | 829 (80%) | | |

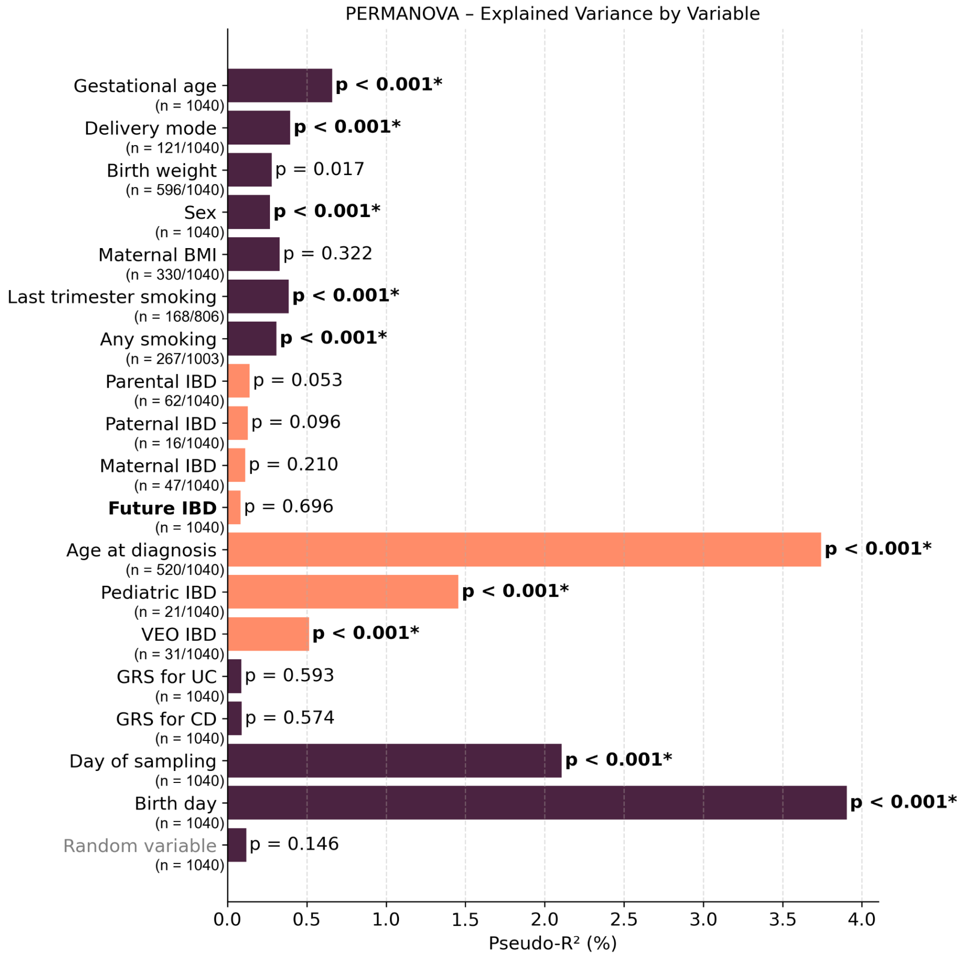


**Figure S2**: Contribution in percentage to the total metabolome variance before correcting for sampling year and postnatal sampling day. Gestational age, delivery mode, and sex are still statistically significant contributors, explaining respectively 0.7, 0.3, and 0.25% of the total variance. Age at diagnosis and VEO are significant, but a proxy of the birth year: the older the sample, the more time the individual had to develop IBD, and vice versa. The signal disappears after correction. Significant p-values are indicated with an asterisk. IBD-related variables are represented in orange. Pseudo-$R^2$ was calculated with PERMANOVA.

### Packages used

The prediction pipeline was implemented in **Python v3.11.13** using the following package versions: pandas 2.2.2, numpy 2.1.2, joblib 1.5.1, xgboost 3.0.5, and scikit-learn 1.7.2.

Variance explained analyses were conducted in **R v4.4.2** using the *adonis2* function in the *vegan* package. The R environment included: dplyr 1.1.4, vegan 2.6.8, stats 4.4.2, glmnet 4.1.8, ggplot2 3.5.1, splines 4.4.2, parallel 4.4.2, pROC 1.18.5, foreach 1.5.2, and doParallel 1.0.17.

All scripts are available at the GitHub repository:
https://github.com/alicefracchia/Neonatal-Metabolome-Prediction-Pipeline
